# Circadian rhythmicity of symptomatic phenotypes in multiple sclerosis: the CircaMS study protocol and feasibility of biomarker collection

**DOI:** 10.1101/2024.10.17.24315625

**Authors:** Doriana Taccardi, Hailey GM Gowdy, Vina Wenyu Li, Ana Cristina Wing, Moogeh Baharnoori, Marcia Finlayson, Nader Ghasemlou

**Affiliations:** Department of Biomedical & Molecular Sciences, Queen’s University, Kingston, ON, Canada; Department of Medicine, Queen’s University, Kingston, ON, Canada; MS Clinic, Neurology, Kingston Health Sciences Centre, Kingston, ON, Canada; School of Rehabilitation Therapy, Queen’s University, Kingston, ON, Canada; Department of Anesthesiology & Perioperative Medicine, Kingston, ON, Canada; Centre for Neuroscience Studies, Queen’s University, Kingston, ON, Canada

**Keywords:** multiple sclerosis, circadian rhythmicity, biomarker, fatigue, pain, mood

## Abstract

**Introduction:** Multiple Sclerosis (MS) is a chronic autoimmune neurological disease with a variable prognosis and unpredictable course. Fatigue, pain, and low mood are common symptoms that tend to fluctuate in people with MS (pwMS). Disrupted circadian rhythms may have a role in the symptoms’ variability. Distinguishing inter-individual differences and temporal daily fluctuations in MS symptoms may help to define specific symptomatic phenotypes. Understanding how these phenotypes are associated with quality-of-life and their immunological underpinnings – immune profiles – could shape new MS management strategies. Our primary aim is to document ongoing fluctuations in fatigue, pain, and mood in a cohort of pwMS to determine whether symptom variability is associated with differential quality of life. Our secondary aim is to evaluate the feasibility of our study design to identify immune profiles of circadian rhythmicity in MS.

**Methods and analysis:** This observational cohort study examines individual temporal fluctuations in MS symptomatology via self-report questionnaires in a cohort of pwMS. All participants complete 1) a baseline battery of questionnaires; and 2) electronic symptom-tracking diaries to rate fatigue, pain intensity, and mood on a 0-10 scale at 3 time-points (08:00, 14:00, 20:00) for 10 days. A subgroup of ∼20 participants – feasibility study – will also complete blood sample collection twice within 24 hours to study immune profiles and molecular markers of circadian rhythmicity in MS. Participants will be grouped into symptomatic phenotypes based on longitudinal data from e-diaries. We will assess whether exhibiting a specific phenotype is associated with certain baseline measures. Flow cytometry, whole blood RNA sequencing, and plasma analyses will be applied to determine changes in immune profiles indicative of circadian rhythmicity.

This work has the potential to reduce the burden of this complex disease on a global scale. Future studies will build on our work to understand individual variability in MS symptomatology, including disease severity; identification of biomarkers underlying the association between rhythmic symptomatology profiles and symptomatic phenotypes in MS; and designing personalized interventions focused on inter-individual differences in symptomatology and circadian rhythmicity.

**Ethics and dissemination:** The CircaMS project and its associated procedures have been reviewed and approved by the Queen’s University Health Sciences and Affiliated Teaching Hospitals Research Ethics Board (File number: 6039383). Participants provide informed consent to participate, and their data will not be identifiable in any publication or report. All documents are stored securely and only accessible by study staff and authorized personnel. Results will be presented to academic and lay audiences via national/international conferences, publications in peer-reviewed journals, social media, and through an official website created to engage pwMS, caregivers, clinicians, and researchers.

**Strengths and limitations of this study:**

- We use electronic symptom diaries to ensure the ecological validity of self-reported baseline questionnaires and temporal variations in fatigue, pain, and mood validated in the MS population.
- This is the first study assessing circadian rhythmicity in multiple sclerosis using a biopsychosocial approach.
- A feasibility study will determine if diverse immune profiles and molecular markers of circadian rhythmicity exist in pwMS.
- Data for the national study will be collected using self-report questionnaires only, which may lead to random or systematic misreporting.
- The online nature of the study might affect the diversity in our sample (eg, the representation of rural and/or underprivileged communities).

## 1. Introduction

Multiple Sclerosis (MS) is a chronic immune-mediated disease of the central nervous system characterized by demyelination and neurodegeneration. This heterogeneous disease has a variable prognosis and unpredictable disease course(1). The prevalence of MS has seen a global increase since 1990, with the highest age-standardized prevalence in high-income regions like North America, Western Europe, and Australasia (2). Fatigue (3), pain (e.g., headache, neuropathic pain, and lower back pain) (4), and low mood (5) are among the most common symptoms reported by people with MS (pwMS). Emerging evidence suggests substantial variability in the occurrence and intensity of MS symptoms such as fatigue (physical and cognitive)(6), pain(7), and low mood(7) and how they are perceived on a moment-by-moment and day-to-day basis. The ecological momentary assessment (EMA) is a valid methodology to characterize temporal variations in self-reported symptoms and identify daily fluctuations in pain, fatigue, and low mood within the MS population (8, 9).

It is necessary to consider the influence of circadian rhythms (or, 24-hour cycles) when investigating variations in processes that have both a psychological and physiological component, such as fatigue, pain, and low mood. Disrupted circadian rhythms can impact MS symptoms and may play a role in their daily fluctuations(10). Circadian rhythms align physiological functions with the environment(11) and are directed by an endogenous clock system, located in the suprachiasmatic nucleus, which controls the release of hormones and other secreted factors. Circadian clocks work in a tightly regulated feedback process lasting ∼24h that influences biomarkers (e.g., gene expression and neuroinflammation) and psychosocial experiences (e.g., fatigue, mood)(10).

Normal fluctuations of clock genes across 24h are significantly reduced in the mouse model of MS (experimental autoimmune encephalomyelitis, EAE), suggesting clock-dependent circadian rhythm disturbances(12). In the clinical population, genetic polymorphisms in key circadian genes are associated with the risk of developing MS in the clinical population(11). Additionally, pwMS possess lower levels of secreted mediators including the circadian hormone melatonin and pro-inflammatory cytokines(13–15). Thus, understanding whether rhythmic expression of immune markers (i.e., at the RNA or protein level) and serum proteins involved in MS impact symptoms like fatigue, pain, and low mood may help identify new therapeutic avenues.

Approved biomarkers for MS (e.g. magnetic resonance imaging, spinal fluid analysis, evoked potentials, etc.) are mostly used for disease diagnosis and detection of neuroinflammation related to disease activity(16). The integration of biomarkers has revolutionized the management of other chronic conditions like cancer, where oncologists have been able to tailor treatment based on patients’ molecular profiles(17). Similarly, we believe it is crucial to identify prognostic biomarkers in MS to improve the management of various MS symptoms and to apply individualized treatments based on molecular profiles and symptomatic phenotypes. Currently, the burden of MS symptoms such as fatigue, pain, and low mood is substantial, and there is an urgent need for more targeted biomarkers that can guide multi-disciplinary symptom management and monitoring.

Our CircaMS study uses a multi-disciplinary approach to investigate the role of circadian rhythmicity in MS. Our focus is on characterizing key symptomatic phenotypes (daily fluctuations in fatigue, pain, and mood) and understanding how they are associated with baseline quality of life measures. Furthermore, we aim to identify immune biomarkers of circadian rhythmicity in the peripheral blood mononuclear cells (PBMCs) of pwMS. This work may help bring us closer to developing innovative personalized treatment strategies, targeting both inter-individual differences in symptomatology and circadian dysfunction at a molecular level. These are important steps toward the primary missions and research priorities set by the Wellness Research Working Group (National MS Society) and the International Progressive MS Alliance(18, 19) that will lead to ultimately reducing the burden that this complex disease has on Canadian and global populations.

## 2. Methods and analysis

This study adheres to the principles of the Declaration of Helsinki(20) and the results will be reported in accordance with STROBE guidelines(21). The CircaMS study and its associated procedures have been reviewed and approved by the Queen’s University Health Sciences and Affiliated Teaching Hospitals Research Ethics Board (File number: 6039383), with the latest approval on May 09, 2024.

### 2.1 Study design and aims

The CircaMS study explores how circadian rhythms and daily fluctuations in fatigue, pain, and mood in pwMS might be associated with quality of life assessed at baseline. Our primary aim (*national/international cohort*) is to document ongoing intra-daily fluctuations in fatigue, pain, and mood (symptomatic phenotypes) in a cohort of pwMS to determine whether symptom variability is associated with specific measures of quality of life. Our secondary aim (*local feasibility study*: sub-cohort of ∼20 people) is to evaluate the feasibility of our study design to identify immune profiles of circadian rhythmicity in pwMS (fig. 1). To address these outcomes, the CircaMS study will recruit pwMS for approximately 4 years collecting data using established methods from epidemiology (surveys and e-diaries) and neuroimmunology (blood samples).

**Figure 1.**
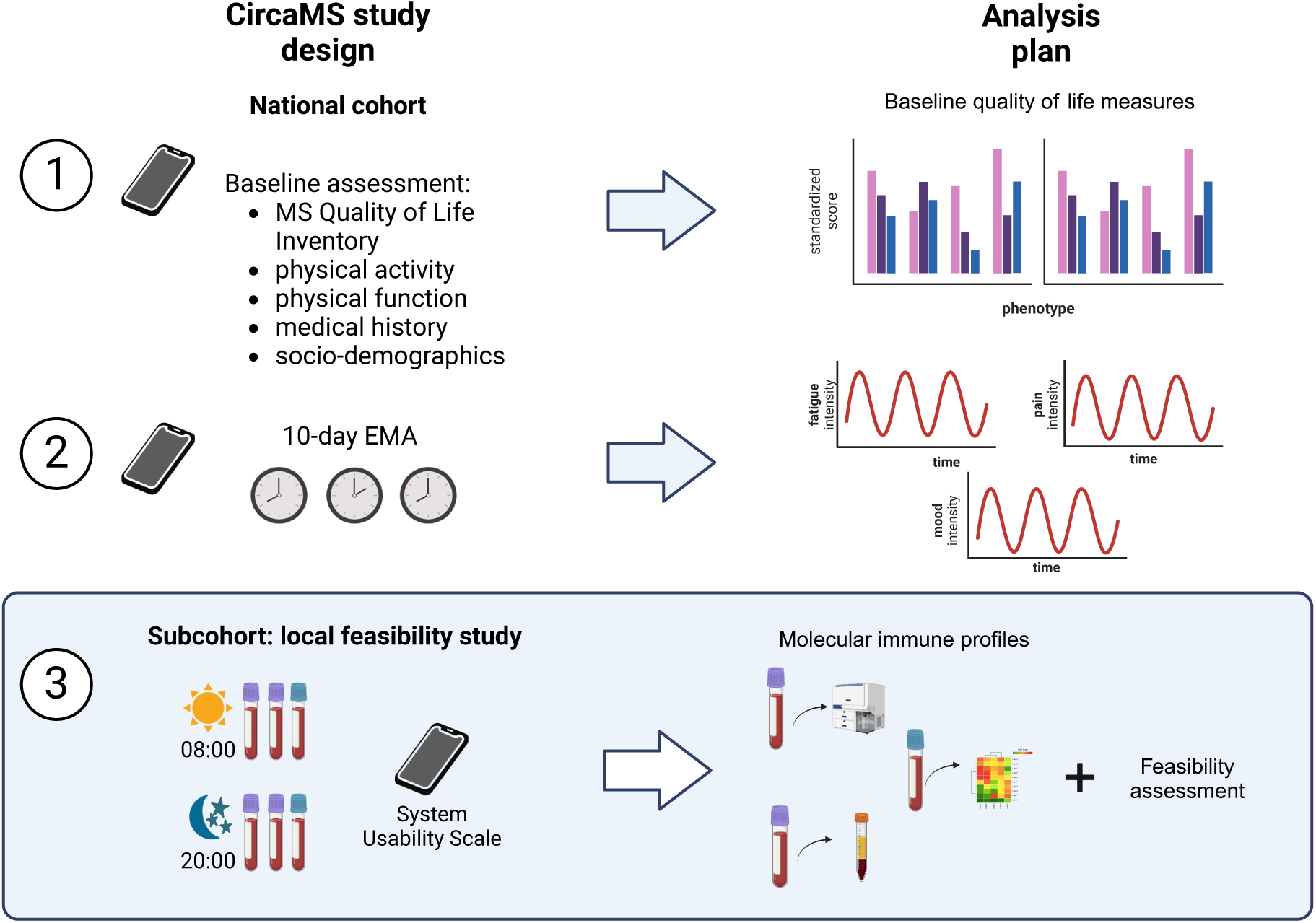
CircaMS Study design: The study is divided into two parts: National cohort consisting of a baseline assessment that includes a battery of questionnaires and a 10-day e-diary. In addition, a sub-cohort of ∼20 people will also complete the collection of biological samples twice a day for flow cytometry, plasma, and RNA sequencing. The feasibility component of the protocol will be assessed in this sub-cohort, recruited in Kingston through the local MS clinic and research centre.

### 2.2 Secondary aim – Local feasibility study

Evaluate the feasibility of our study design in pwMS, including survey completion and blood sample collection to study the circadian expression of inflammatory markers/immune profiles of circadian rhythmicity. To assess the feasibility of our study procedures for the local study (22), we have set up four sub-aims:

1. *Recruitment capability:*

- *Can we recruit 20 people with MS within 18 months for both surveys and 2 blood samples collected within 12 hours?*
- *Are the eligibility criteria suitable or too inclusive or restrictive?*
- *What are the reasons for ineligibility or refusal?*
2. *Data collection procedures*

- *Are data collection procedures (i.e., repeated venipuncture: blood sample collection) manageable for the MS population?*
- *Are the planned EMA e-diary times manageable for participants?*
- *Are the planned times for blood collection convenient for participants or does it need to be changed?*
3. *Acceptability of the procedures*

- *Can fidelity to the protocol be achieved?*
- *Are there any unexpected adverse events involving both participants and the research team?*
- *Are the procedures/measures used acceptable and usable?*
4. *Proof of concept*

- *Are there any changes in the molecular profiles of participants as expected?*
- *Are the preliminary findings congruent with the hypothesis?*
- *If there are no changes, are the collection procedures appropriate?*

### 2.3 Eligibility criteria – National/international cohort and Local feasibility study

Participants who are at least 18 years of age, have internet access (any device is accepted), are fluent in English or French (or additional languages, as the study is translated), and have subjective complaints of fatigue OR pain are eligible to participate in CircaMS; participants with mood complaints alone (e.g. no fatigue or pain) will not be eligible. Participants are excluded if they have any other neurological condition (e.g., stroke, dementia), severe and untreated psychotic disorder (e.g., schizophrenia), diagnosis of an untreated and severe primary sleep disorder such as narcolepsy (sleep apnea is accepted), and/or history of travel across time zones in the past three weeks or are planning to travel to a different time zone during the period of data collection.

### 2.4 Recruitment and participants – National/international cohort

The study is open nationally and internationally for collection of epidemiological data (surveys) to reach >200 participants with complete data in approximately 2 years. These numbers are expected to be sufficient to group participants in meaningful symptomatic phenotypes, based on our previous unpublished results on a population of people with chronic low back pain where we found four major pain phenotypes (constant low, constant high, rhythmic, and mixed patterns) at an approximate ratio of ∼1:1:1:1. The sample size calculation was based on ANOVA assuming a type I error *α* = 0.05, and type II error *β* = 0.2 (i.e. 80% power). To detect a difference in pain, fatigue, and mood across phenotypes, we will require around ∼50 participants per pain phenotype. The study is advertised via the research team’s official social media pages (Facebook, X, etc.), a dedicated study website <u>CircaMS</u>, and paper forms (e.g., posters, handouts). We also leverage MS networks, people with lived experience (PWLEs) and collaborators to help with recruitment.

### 2.5 Recruitment and participants – Local feasibility study

The feasibility study will recruit at least 20 adults from Kingston Health Science Centre (KHSC) MS clinics within 2 years of study recruitment initiation, with a clinician’s diagnosis. Based on our previously described unpublished study, detecting differences in circadian genes will require ∼20 participants per phenotype. We will start collecting blood from 20 pwMS to assess the feasibility of these procedures before expanding to a larger sample sufficiently powered for this aim. Participants for the local study are recruited primarily through the KHSC MS clinic by clinicians and researchers. Additionally, recruitment material will be sent to other clinics and local services accessed by pwMS. Individuals have the freedom to follow the link to initiate study enrolment (eligibility screening questions) through the REDCap platform(23, 24). Participants can opt in or out of the blood collection using the REDCap link.

### 2.6 Theoretical framework and considerations for the analyses

The National Institute of Health established a symptoms science model to help guide precision medicine and develop innovative interventions(25). According to this model, it is important to consider the presentation of different symptoms (e.g., psychological and physiological comorbidities) to determine their phenotypic characteristics and to test biomarkers (e.g., genes, protein)(25, 26). This allows clinicians/researchers to consider several domains that interact with symptom experiences, such as personal characteristics, functional outcomes, health/illness factors, environment and management strategies(27). When Patient-Reported Outcomes in electronic form(28) are employed to capture data, using validated and standardized measures (e.g. PROMIS(29)), it becomes possible to include mental, physical and social well-being components in the model(30). This is particularly important when studying chronic and multifactorial diseases like MS; where the combination of clinical profiling, real-world data collection, and biomarker identification is key for a comprehensive understanding of individual differences within the disease(31).

The CircaMS baseline measures include several standardized questionnaires to account for quality of life in MS (e.g. Multiple Sclerosis Quality of Life Inventory (MSQLI)(32), PROMIS Fatigue MS-8a(33)) (table 1). We will include those variables as covariates in our regression model and our final modelling will be based on all these considerations. Ultimately, this together with phenotyping based on the EMA e-diary measures and biomarker identification via blood sample will establish a solid model to study MS, with the vision of fostering precision medicine and personalized treatment.

**Table 1.**
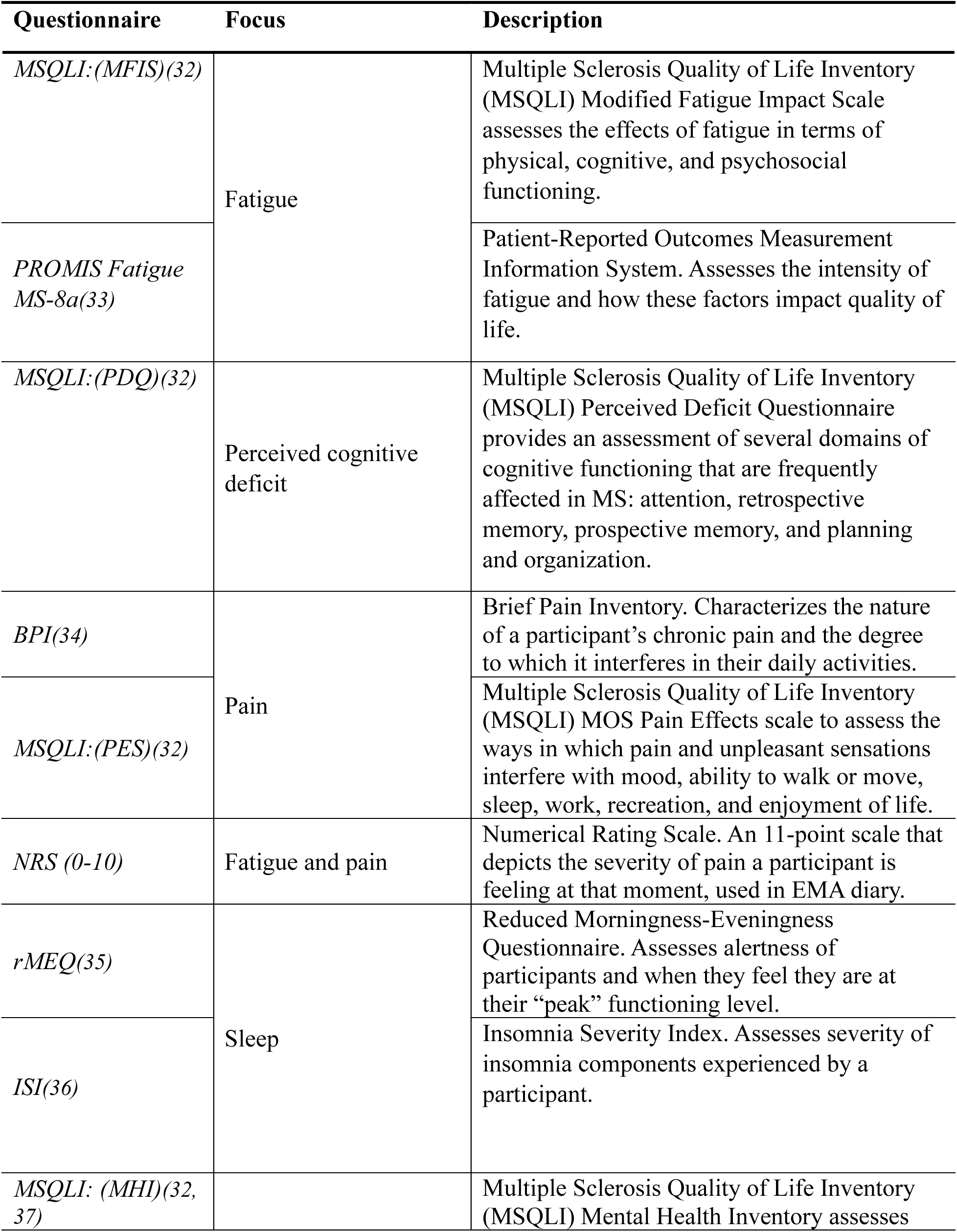
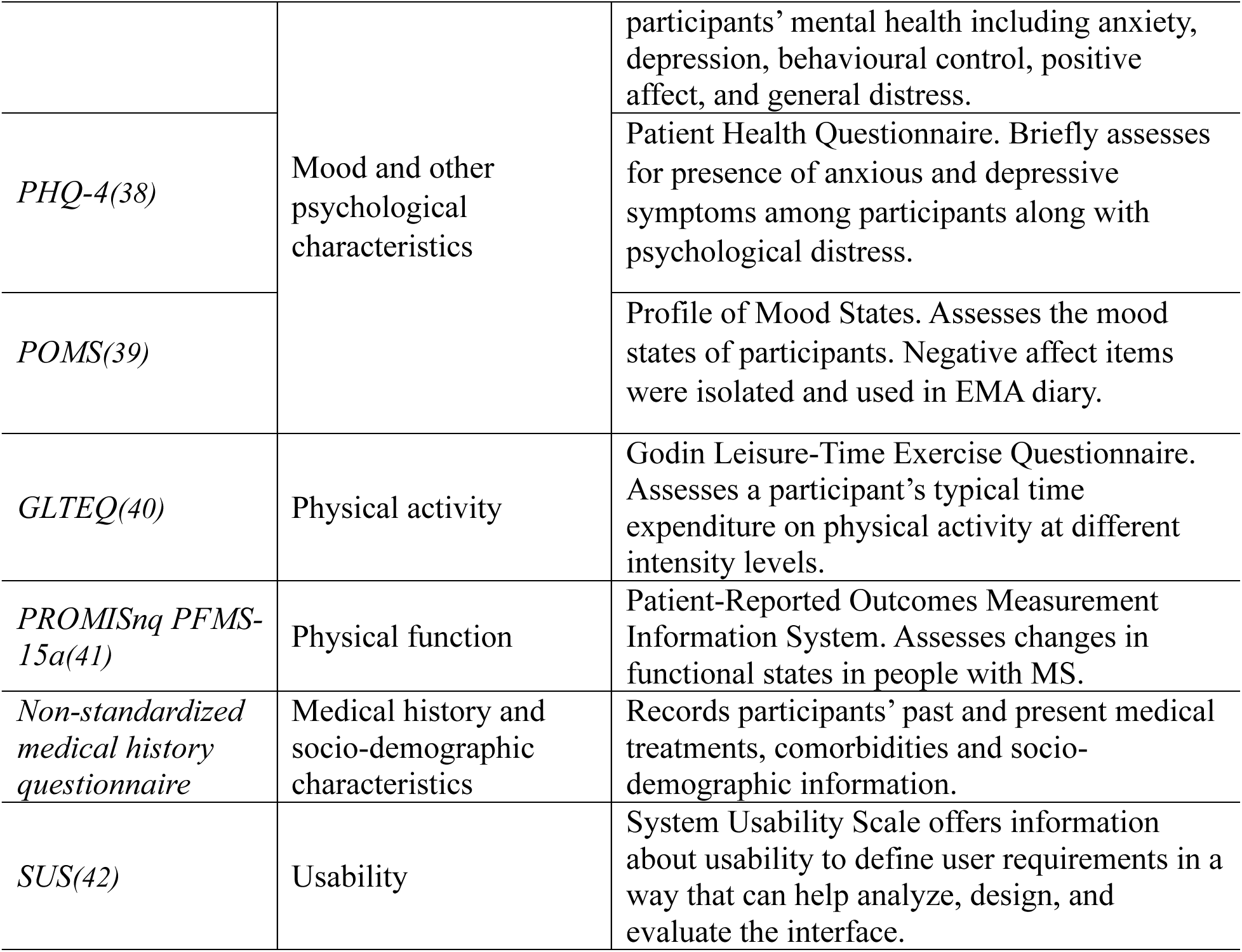
Validated and non-standardized measures completed by participants in the CircaMS baseline battery and EMA symptom diaries.

### 2.7 Data collection – National/international cohort

Eligible pwMS complete an online form with a link to a REDCap survey for informed consent, and baseline questionnaires, followed by our e-diary using the EMA approach(8).

*Baseline questionnaire battery* includes self-report questionnaires covering different domains: fatigue, pain, sleep, mood and other quality of life variables, physical activity, physical function, and medical history and socio-demographic characteristics (table 1).

### 2.8 10-day e-diary (EMA)

following baseline assessment, all participants are asked to rate their fatigue, pain, current mood, and activity levels using a Numerical Rating Scale (NRS) scale three times a day for 10 days using a 10-day e-diary. Participants complete the e-diary 3 times daily (8:00, 14:00, 20:00) for 10 days, providing an email address or phone number to receive a personalized link tracking their daily reports via REDCap/Twilio Integration Module. An additional question about hours of sleep and perceived sleep quality is included in the 8:00 survey. Invites are sent according to the local time where the participant is based. Surveys completed past 10:00, 16:00, 22:00, are excluded from the analysis as timed-out (no longer ecologically valid). Any symptom diary containing ≥5 days with submitted diaries at all time points over the 10 days will be used in the analyses; all other symptom diaries are excluded from main analyses and may be used for secondary analyses.

### 2.9 Data collection – Local feasibility study

PwMS participating in the local feasibility study complete all the measures mentioned in the previous section. In addition, MS clinicians/researchers will record information relevant to the study (such as Vitamin D levels or medication use) using electronic medical records.

*Blood collection*: Blood draws are collected after baseline assessment and around the time the last day of EMA has been completed. Participants will be recruited through the MS Clinic. They will be given the choice of either attending the Research facility in Kingston twice daily or receiving a home visit by a certified phlebotomist in the research team if they live in the Kingston area. They will provide 2 blood samples (≤20ml/draw) within a 12h period (morning [7:00 -10:00] and evening [19:00 -22:00] in one single 24-hour cycle. Blood is collected in Tempus or PAXgene® tubes for whole blood RNA sequencing and K2EDTA tubes for flow cytometry and serum proteins. Samples are processed and/or stored as described in sample processing and storage section of this paper.

Twenty participants who completed both blood samples and surveys are sent an additional survey following the last diary completion (10 days) for their assessment of our local study procedures (feasibility sub-aim 3) and the usability of our EMA diary based on the System Usability Scale(42).

### 2.10 Analyses – National/international cohort and local feasibility study

To accomplish our primary aim, participants are grouped by the mean and standard deviation (SD) of their reported EMA pain scores across all time points available using the SD 50^th^ percentile to determine low/high variability groups.(43) When the full sample size is reached, participants with distinct rhythmicity trajectories (i.e., fatigue, pain, mood phenotypes) are identified by EMA scores using a latent class mixed effect model (LCMM, based on *lcmm* R package(44)), a probabilistic modelling algorithm approach that clusters longitudinal data accounting for correlation between repeated measures(45, 46) and has been used to characterize pain chronicity over months(47). To ensure biologically meaningful phenotypes are identified, functional data analysis with high-dimensional data clustering (based on *funHDDC R* package(48)) will be considered in addition to LCMM as an alternative approach; this allows for participants with similar pain fluctuation phenotypes to be clustered together. Graphical tools (e.g., Sankey plots) will be used to visualize changes in phenotype. Furthermore, we will use linear mixed-effect models(49, 50) to test for differences in average pain scores in the morning vs evening across time points. We will use descriptive statistics to characterize the sample and inferential statistics (e.g. Fisher’s Exact tests, chi-square, Kruskal-Wallis, multinomial logistic regression and other regression models) to determine the association between symptomatic phenotypes and baseline well-being/quality of life measures, including possible confounders. All statistical analyses will be performed halfway through and at the end of the study recruitment using R(51) or SPSS(52) where appropriate (fig.1).

### 2.11 Analyses – Local feasibility study

Our secondary aim (feasibility of the procedures) both self-report measures and blood collection and specific sub-aims are assessed as follows:

*1. Recruitment capability:*

- *Can we recruit 20 people with MS within 18 months for both surveys and 2 blood samples collected within 12 hours?*

This pilot study will help us to answer the question of being able to recruit this number of people for 2 blood samples within 12 hours (yes or no). The recruitment rate is also measured by the number of people enrolled over the number of participants approached plus screened plus inquired (these numbers are recorded using enrolment and recruitment logs).

- *Are the eligibility criteria suitable or too inclusive or restrictive?*

We assess the suitability of eligibility criteria by counting the number of people who entered the study but were deemed ineligible following initial screening as recorded in the REDCap eligibility form. Eligibility criteria will be considered suitable if more than 50% of people who entered the study are deemed eligible to take part in the study. If one specific criterion results in the non-eligibility of 50% of participants, that criteria will be considered restrictive and therefore might be adapted.

- *What are the reasons for ineligibility or refusal?*

Recurrent themes/reasons for declining are recorded whenever participants are willing to share them. These will be considered to guide changes in recruitment procedures.

*2. Data collection procedures*

- *Are data collection procedures (i.e., repeated venipuncture: blood sample collection) manageable for the MS population?*

The feasibility of repeated blood sample collection in the MS population is evaluated by counting the number of people who were eligible for the local study but declined the blood sample collection.

- *Are the planned EMA e-diary times manageable for participants?*

The appropriateness of EMA times is assessed by considering the number of missed time points for morning, afternoon, and evening diary completions. Less than 50% completion per time point in more than 50% of the enrolled participants will be considered unmanageable.

- *Are the planned times for blood collection convenient for participants or does it need to be changed?*

Appropriateness of blood collection times is assessed by considering the number of missed time points for morning and evening blood collection sessions. Furthermore, the location of blood collection is evaluated based on the number of appointments scheduled at this location. Alternative locations (e.g. home collection) may be proposed to facilitate recruitment.

Compliance is assessed through examination of the percentage of people who completed ≥5 days with submitted diaries at all time points over the 10-day period and who completed 2 blood draws. Completion rate is evaluated by dividing the number of people who completed the survey by the total number who started the survey without completing it.

*3. Acceptability of the procedures*

- *Can fidelity to the protocol be achieved?*

Adherence to the protocol during data collection is evaluated. The number of deviations from the protocol will be recorded by members of the research team and PI. No more than five deviations from the protocol for every 20 participants will be considered acceptable.

- *Are there any unexpected adverse events involving both participants and the research team?*

The safety of procedures for researchers and participants is estimated by recording the number of adverse events related to blood collection procedures.

- *Are the procedures/measures used acceptable and usable?*

Scores and responses to the System Usability Scale(42) are used to determine the acceptability and usability of our study procedures.

*4. Proof of concept*

- *Are there any changes in the molecular profiles of participants as expected?*

Blood samples will be processed in batches based on collection to determine whether there are any changes in the immune profiles of participants.

- *Are the preliminary findings congruent with the hypothesis?*

Processing and analyses of blood samples will be performed immediately for flow cytometry and after study recruitment (20 pwMS) for RNA sequencing and plasma to avoid batch effects. This will allow us to explore whether the results are congruent with the hypothesis or whether the procedures need to be changed.

- *If there are no changes, are the collection procedures appropriate?*

Statistical analyses will be performed halfway through the study recruitment to explore whether the results are congruent with the hypothesis or whether the procedures need to be changed.

### 2.12 Sample Processing and Storage – Local feasibility study

*Flow cytometry:* Up to 6 ml of blood is used to characterize circadian fluctuations in the immune cell population using cell surface markers. Immune cells are phenotyped as following: CD4+ T cells (CD45+CD3+CD4+CD8-), CD8+ T cells (CD45+CD3+CD4-CD8+), B cells (CD45+CD3-CD19+), neutrophils (CD45+CD11b+CD16+CD14low/int), monocytes (CD45+CD11b+CD16+/- CD14+) and macrophages (CD45+CD11b+CD16+CD14hi). The markers HLA-DR and HLA-ABC are used to identify specific activation states.

*Whole blood RNA sequencing and bioinformatic analysis:* Up to 3 ml of blood is used for whole blood RNA sequencing. All samples are stored at -80°C and/or in liquid nitrogen and sent to a genome facility for simultaneous processing and sequencing to avoid batch-to-batch variability. Purified RNA samples from PBMCs will be used to assess circadian oscillations in global gene expression patterns using differential gene expression, network analysis, and other bioinformatic tools.

*Plasma:* Up to 6 ml of blood is used to collect plasma through centrifugation. The plasma is aliquoted and stored at -80°C and/or in liquid nitrogen. Plasma samples will be processed using multi-omics technologies to profile circadian changes in mediator expression (e.g., proteins, lipids). While serum analysis remains outside the scope of this study, it will be banked for future analysis (fig.1).

### 2.13 Patient and Public Involvement

All research questions and outcome measures were developed in collaboration with people with lived experience of MS, taking into account their priorities and experiences with the disease. This was done through consultations with the research team, they completed the survey and returned feedback about the questions. This guided the final version of our CircaMS measures. CircaMS also has a patient partner as part of the study team, who contributes to the knowledge translation and dissemination efforts by helping the research team summarise the research objectives and background in clear user-friendly language and ensure that the information is accessible to a public audience.

## 3. Ethics and dissemination

The CircaMS project and its associated procedures have been reviewed and approved by the Queen’s University Health Sciences and Affiliated Teaching Hospitals Research Ethics Board (File number: 6039383), with the latest approval on May 09, 2024. The research team will ensure that the participants’ confidentiality is maintained. The participants are identified only by a study participant ID on any electronic database. All documents are stored securely and only accessible by study staff and authorized personnel. The study complies with The Personal Information Protection and Electronic Documents Act (PIPEDA) and the Tri-Council Policy Statement: Ethical Conduct for Research Involving Humans (TCPS).

All information is stored in REDCap(23, 24) online data collection software, or a secure file password protected. Medical records are accessed to gather information relevant to the study and only by an authorized member of the research team with a valid research appointment. Since some of the in-person testing sessions occur outside of normal working hours, a second member of the research team may be present on-site to maintain both participant safety and that of the researchers.

Participants will not be identified in any publication or reports, nor will information such as email addresses appear in any study documents. REDCap has specialized functionality which allows only select users to view and input identifying information, with all other users seeing exclusively de-identified data and prohibiting the downloading of identifying information when exporting the dataset. The study team will present results from this study to research, clinical, and lay audiences via national and international conferences, publications in peer-reviewed journals, and ongoing knowledge translation efforts (e.g., blog, optional return of results to participants) spearheaded by investigators at the lead site. The authors will acknowledge eventual funding and other contributors. The official website <u>CircaMS</u> is used to post updated resources (blogs, videos, research updates, etc.) to engage people who live with MS and their families or friends, clinicians or anyone interested in the topic.

## 4. Expected results and discussion

We expect that people with rhythmic EMA symptomatic phenotypes (based on fatigue and pain daily fluctuations) will report better quality of life measures and healthier molecular profiles through examination of gene expression patterns and PBMC activation states, compared to people with constant EMA symptomatic phenotypes. Our short-term goal is to examine the feasibility of this study design in the MS population and identify circadian rhythmicity as a biomarker for fatigue/pain/mood phenotypes. This will respond to the immediate need 1) to find biomarkers to monitor and prognosticate common MS symptoms, 2) to develop effective symptom management strategies by targeting circadian rhythms; important steps toward the primary missions and research priorities set by the Wellness Research Working Group(18, 19) to reduce the burden that this complex disease has on the global population.

Our long-term goal is to build the foundation to develop new treatment strategies targeting circadian dysfunction. Non-pharmacologic means can include light therapy to reset circadian clocks or chronotherapy to time medication dosing according to symptom need; pharmacologic strategies can be used to target clock gene expression or PBMC activation states at a molecular level. This work will help reduce the burden this complex disease has on the global population. Future studies will build on our work to understand individual variability in MS symptomatology, including disease severity; identification of biomarkers underlying the association between rhythmic symptomatology profiles and symptomatic phenotypes in MS; and designing personalized interventions focused on inter-individual differences in symptomatology and circadian rhythmicity.

Our study examines inter-individual and temporal variations in fatigue, pain, and mood in people with MS, using electronic symptom diaries to ensure the ecological validity of self-reported symptoms and baseline questionnaires validated in the MS population(7, 9). Collecting blood two times per day in our local feasibility study will allow us to understand whether immune profiles and molecular markers of circadian rhythmicity exist in pwMS. Some limitations of our study design include the fact that no formal medical validation of participants’ MS diagnosis is included in the epidemiological portion of the study as all measures are self-reported. For our local feasibility study, a medical validation is instead required. Furthermore, participation may be difficult for individuals with limited computer skills as the study is available online through a web-based platform. However, for our local feasibility study, researchers can offer alternatives to complete the intake survey and support participants in setting up reminders to complete e-diaries.

## Data Availability

Not applicable, this is a study protocol

## Contributors

**Doriana Taccardi:** Conceptualization, Methodology, Software, Writing – Original Draft, Writing – Review & Editing, Project administration. **Hailey GM Gowdy:** Methodology, Software, Writing – Review & Editing, **Vina Wenyu Li:** Methodology, Writing – Review & Editing, **Ana Cristina Wing:** Methodology, Writing – Review & Editing, **Moogeh Baharnoori:** Conceptualization, Methodology, Writing – Review & Editing, Funding acquisition. **Marcia Finlayson:** Conceptualization, Methodology, Writing – Review & Editing, Supervision, Funding acquisition. **Nader Ghasemlou:** Conceptualization, Methodology, Writing – Review & Editing, Supervision, Funding acquisition.

## Funding

This work was supported by the Queen’s University Faculty of Health Sciences Botterell-Howe- Powell Fund (to MB, MF, and NG).

MS Canada supports this work with an endMS Doctoral Studentship Award to DT.

## Competing interests

None of the authors has competing interests related to this manuscript.

## Notes

### Competing Interest Statement

The authors have declared no competing interest.

### Funding Statement

This work was supported by the Queens University Faculty of Health Sciences Botterell-HowePowell Fund (to MB, MF, and NG).
MS Canada supports this work with an endMS Doctoral Studentship Award to DT.

### Author Declarations

The CircaMS project and its associated procedures have been reviewed and approved by the Queens University Health Sciences and Affiliated Teaching Hospitals Research Ethics Board (File number: 6039383).

